# Prevalence of Obesity According to Body Mass Index, Waist Circumference, and Waist-to-Height Ratio in Peru: A Systematic Review and Meta-Analysis

**DOI:** 10.1101/2024.05.21.24307695

**Authors:** Luisa Erika Milagros Vásquez-Romero, Fiorella E. Zuzunaga-Montoya, Joan A. Loayza-Castro, Enrique Vigil-Ventura, Willy Ramos, Víctor Juan Vera-Ponce

**Author notes:** Correspondencia: Vera-Ponce. Luisa Erika Milagros Vásquez Romero, Fiorella E. Zuzunaga-Montoya, Joan A. Loayza-Castro, Enrique Vigil-Ventura, Willy Ramos, Víctor Juan.

## Abstract

**Introduction:** Obesity is a global public health issue with significant health implications. However, studies on the prevalence of obesity in Peru have yielded varied results, highlighting the need for updated data to inform effective public health policies.

**Objective:** The primary objective of this study is to determine the prevalence of obesity in Peru using three anthropometric measures: body mass index (BMI), waist circumference (WC), and waist-to-height ratio (WHtR).

**Methods:** Between March and April 2024, a systematic review of published studies reporting the prevalence of obesity in Peru was conducted. The databases Scopus, Web of Science, Embase, PubMed, LILACS, and Scielo were Searched.

**Results:** Overall, the prevalence of obesity was 23.23%, 38.90%, and 81.53% according to BMI, WC, and WHtR, respectively. However, these figures show wide variability, ranging from 13.10% to 37.4% according to BMI and from 19.4% to 51.6% according to WC. The highest reported prevalence of obesity by WHtR was 85.4%. Nonetheless, only a fraction of these studies were published in the last five years, and few specifically focused on obesity as the primary objective.

**Conclusions:** The prevalence of obesity in Peru varies significantly depending on the anthropometric measure used. To improve the collection and frequency of data on obesity in Peru, it is recommended that cut-off points be standardized to be suitable for the country and that annual national surveys specifically designed for this purpose be implemented.

## Introduction

Obesity is among the most urgent health issues worldwide. It raises risks of metabolic diseases, risks death rates, and leads to many other health complications ^(1)^. Projections say by 2025, one in five adults will face obesity globally. Regions like Southeast Asia and Africa are expected to see big jumps in these numbers. Economically, this issue accounts for about 13% of all healthcare costs—almost $990 million yearly ^(2)^.

In Latin America, roughly a quarter of people struggle with obesity; the trend keeps climbing despite global changes. Data from the Latin American Study of Nutrition and Health (ELANS, an acronym in Spanish) shows that out of 9,218 participants, around 25.2% were obese. In Peru specifically, this study found an obesity rate of 22%, measured BMI as a measure ^(3)^.

Obesity rates in Peru vary according to national surveys. The National Household Survey (ENAHO) from 2012-2013 indicated an obesity prevalence of 19.7% using BMI ^(4)^. Additionally, the Food and Nutrition Surveillance by Life Stage Adult Survey (VIANEV, acronym in Spanish) from 2017-2018 showed higher numbers: 26.8% with BMI, 50.4% with waist circumference (WC), and up to 85.4% with waist-to-height ratio (WHtR) ^(5)^. A more recent study by Vásquez-Romero utilizing data from the National Demographic and Health Survey (ENDES, acronym in Spanish) showed national obesity rates of 25.65%, 42.04%, and 46.49%, based on BMI, WC, and WHtR, respectively ^(6)^.

These differing results highlight the need to investigate how obesity prevalence differs according to different measures at a national level; hence, this study aims to find out how common obesity is in Peru by looking at three measures—BMI, WHtR, and WC—to give a comprehensive perspective of the epidemic within Peru context.

## Methodology

### Study Design

This study is a systematic review and meta-analysis of the prevalence of obesity in Peru. The PRISMA 2020 statement was used to ensure the proper conduct of systematic reviews and meta-analyses ^(7)^; however, it was adapted considering this was a different type of systematic review from the classic ones recommended by this guideline.

### Search Strategy

Between March 1 and April 1, 2024, a systematic search was conducted in Scopus, Web of Science, Embase, PubMed, LILACS, and Scielo databases. The key terms used included “prevalence,” “obesity,” “body mass index,” “abdominal circumference,” and “waist-to-height ratio.” The complete search strategy is available in Supplementary Material 1.

### Selection Criteria

The inclusion of studies was evaluated based on the following criteria: 1) original articles, 2) those employing probabilistic sampling techniques, which are essential to ensure that the results are generalizable to the study population, and 3) those that assessed obesity using at least one of the following anthropometric parameters: BMI, WC, or WHtR. Conversely, studies focused on selective populations, such as groups with specific health conditions, were excluded as these could introduce biases in evaluating obesity prevalence.

### Study Selection

With the search results, the first phase involved the removal of duplicates using the Rayyan software for article storage. Abstracts of articles/conferences and unavailable manuscripts were excluded.

In the second phase, two investigators (FEZM and JLC) independently reviewed the titles and abstracts of the manuscripts. If there was agreement on inclusion, the manuscript was added; otherwise, it was excluded. In case of any discrepancy or doubt about inclusion, a third author (LEMVR) resolved it.

Finally, the full text of the included manuscripts was verified and added to an Excel sheet. The investigators also carried out this process, and the fourth author resolved any doubts.

### Data Extraction and Qualitative Analysis

Using Microsoft Excel 2016, the following specific data were extracted from each study: first author and year, city, inclusion criteria, exclusion criteria, sample size, sex (% female), age, and diagnosis of obesity according to BMI, WC, and WHtR.

### Risk of Bias Assessment

Munn Z et al. ^(8)^ assessed the quality of the selected articles using the risk of bias tool for prevalence studies. This tool consists of nine questions with three standard response options (include/exclude/seek more information) based on the evaluator’s criteria.

The study’s risk of bias was evaluated based on the following nine criteria: 1) Was the sample frame appropriate to address the target population? 2) Were the study participants recruited appropriately? 3) Was the sample size adequate? 4) Were the study subjects and setting described in detail? 5) Was the data analysis conducted with sufficient coverage of the identified sample? 6) Were valid methods used for the identification of the condition? 7) Was the condition measured in a standard and reliable manner for all participants? 8) Was the statistical analysis appropriate? 9) Was the response rate adequate, and if not, was the low response rate managed appropriately?

The level of bias is assessed by calculating the total number of criteria with an affirmative response and converting this score into a percentage (n/9). Studies scoring below 50% are considered high risk of bias, 50-69% moderate risk of bias, and 70% and above low risk of bias. The quality assessment tool was initially tested on a small number of studies. Two additional investigators were available for consultations and to resolve any disagreements.

### Statistical Analysis

All quantitative analyses were performed using STATA version 18 (Stata Corp, College Station, TX, USA). All sources providing data on BMI, WC, and WHtR prevalence were included in the meta-analysis and pooled analysis. The total sample size and the number of cases for each condition were extracted to calculate prevalences and their 95% confidence intervals (95% CI).

A random-effects model was used to combine the prevalences, as significant heterogeneity was expected between the studies due to differences in study populations, measurement methods, and other factors. Heterogeneity among the studies was assessed using the I² statistic. The interpretation of heterogeneity followed the Cochrane Handbook guidelines: 0 to 40% = might not be important; 30 to 60% = moderate heterogeneity; 50 to 90% = substantial heterogeneity; 75 to 100% = considerable heterogeneity.

Sensitivity analyses were performed to assess the robustness of the results by excluding studies with a high risk of bias or different measurement methods. Additionally, sensitivity analyses by year (differentiating between < 2014 and ≥ 2013), by sex, and by scope (subnational vs. national) were conducted.

### Ethical Considerations

This study is based on analyzing manuscripts published in journals and scientific databases, so the risks involved are minimal. Therefore, the authors did not consider it necessary to submit the project to a research ethics committee.

## Results

### Study Selection

A total of 4,529 publications were found, and after removing duplicates, 3,094 unique articles remained. From these, 1,435 manuscripts were evaluated, of which 70 articles met the inclusion criteria. Finally, 17 manuscripts were included in the analysis. Details are provided in Figure 1.

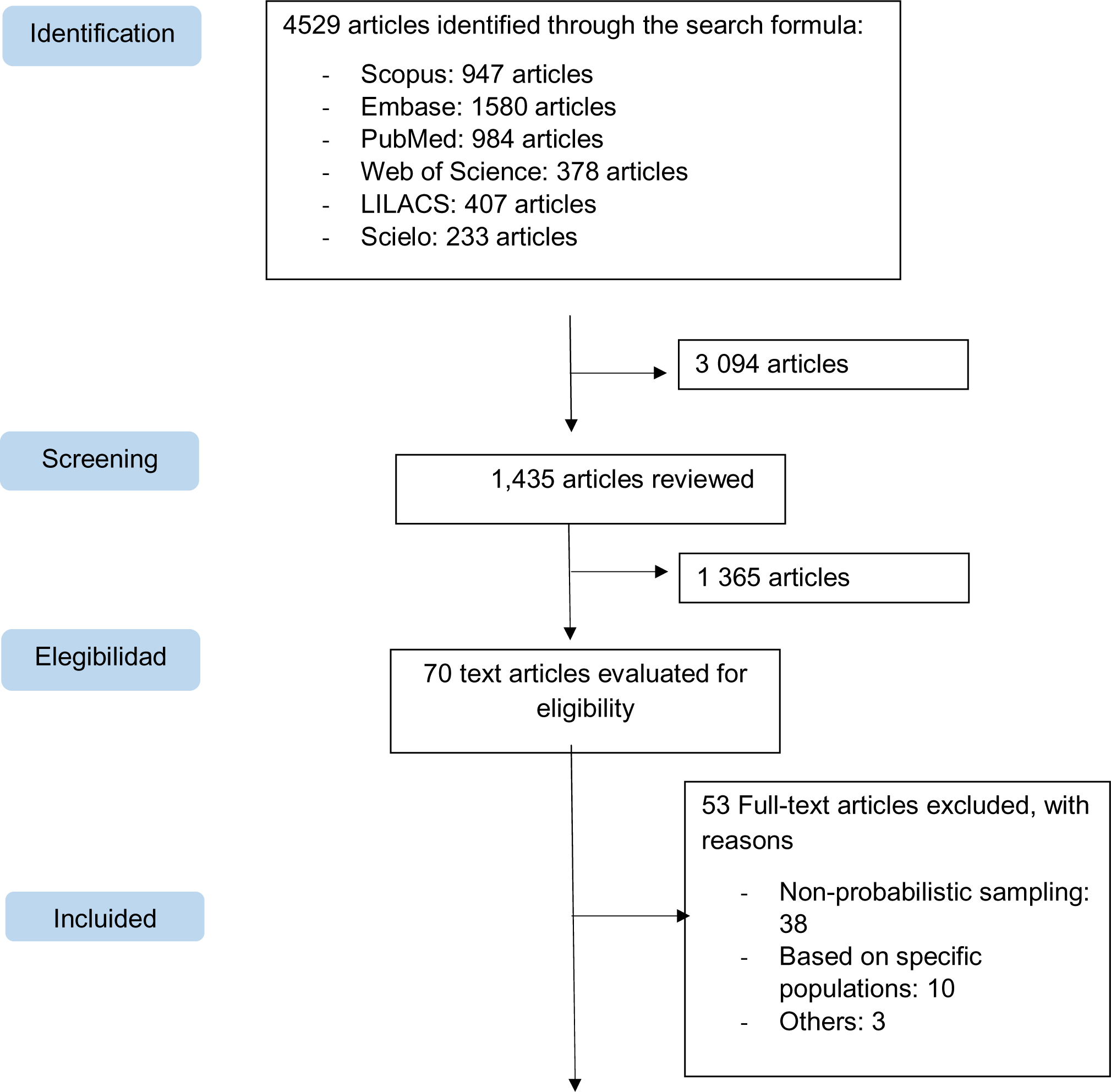
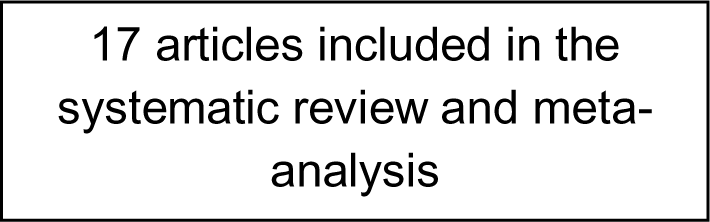

### Study Characteristics

The studies, ranging in date from Jacoby (2003) ^(9)^ to Vásquez-Romero (2024) ^(6)^, presented sample sizes from 217 ^(10)^ to 35,255 individuals ^(6)^. Four studies were national in scope, while 13 were subnational. Of the total, 16 studies evaluated obesity by BMI, 12 by WC, and 5 by WHtR.

A variety of approaches were observed regarding the databases used. Some studies developed and used databases tailored to their specific research needs. For example, studies by Seclen ^(11)^, Herrera-Enriquez ^(10)^, Adams and Chirinos ^(12)^, and Barboza Palomino ^(13)^ designed their datasets to study obesity in specific populations in Lima, Arequipa, and Huancayo. Conversely, several studies utilized existing databases from previous research, offering an established framework for longitudinal or comparative analysis. The National Survey of Indicators for Non-Communicable Chronic Diseases (ENINBSC) ^(14)^ collected diet, exercise, and other anthropometric indicators data. Medina-Lezama ^(15)^ used data from the PREVENTION study, while Revilla L. ^(16)^ employed data from the FRENT LIMA-CALLAO study, and Carrillo-Larco et al ^(17)^ used data from the CRONICAS study. Additionally, some studies used national survey databases such as ENAHO ^(4)^, VIANEV ^(5)^, and ENDES, which provide extensive and representative information on the Peruvian population. These surveys are particularly valuable for epidemiological studies as they include a wide range of data on health, nutrition, and socioeconomic factors at the national level.

Significant variations were observed in the analysis of obesity prevalence across different studies. The study by Adams and Chirinos reported the highest prevalence for both BMI (37.40%) and WC (51.6%) ^(12)^. In contrast, the ENINBSC study ^(14)^ showed the lowest prevalence for BMI at 13.10%, and Jacoby ^(9)^ recorded the lowest for WC at 19.4% ^(9)^. Regarding WHtR, Aparco and Cárdenas-Quintana ^(5)^ reported the highest prevalence at 85.40%, while Vásquez-Romero ^(6)^ reported two prevalences for WHtR depending on the cut-off point: ≥ 0.50 (84.78%) and ≥ 0.59 (48.05%).

The percentage of female participants in the reviewed studies varied considerably, with the lowest being 50.13% (Adams and Chirinos) ^(12)^ and the highest at 71.1% (ENINBSC) ^(14)^, which could influence the prevalence variations observed between studies. Regarding the geographical distribution of the studies, there was a higher concentration of research in urban areas, particularly in the capital, Lima, followed by major cities such as Tumbes and Arequipa.

Among studies that included younger populations, Revilla L ^(16)^. included individuals aged 15, standing out with the lowest age limit among all listed. On the other hand, studies with a broader age range, with an upper limit of 99 years, include those by Vásquez-Romero ^(6)^, ENINBSC ^(14)^, and Seclen ^(11)^, covering a considerable diversity in adult and elderly ages.

During this systematic review, a limitation was encountered in not identifying manuscripts published in indexed databases that met all proposed objectives. However, aware of the importance of a comprehensive approach, we resorted to non-indexed national data sources to complement scientific publications. Specifically, we accessed and extracted pertinent data from ENINBSC ^(14)^, which, despite not being indexed, provides vital information for our research. Similarly, we analyzed the PERU MIGRANT study database ^(18)^, using the study by Miranda JJ ^(19)^. as a reference. Although the associated publication did not report obesity prevalence data by WC and WHtR, we extracted these data directly from their database. This strategy allowed us to include a broader range of data and provide a more robust and representative analysis of obesity prevalence in Peru, thereby maximizing the utility and impact of our findings in understanding this public health issue.

### Meta-Analysis on Obesity Prevalence

According to BMI, the pooled prevalence of obesity was 23.23% (95% CI: 20.00%— 26.63%), but the heterogeneity was quite high (I² = 99.2%) (Figure 2).

**Figure 2.**
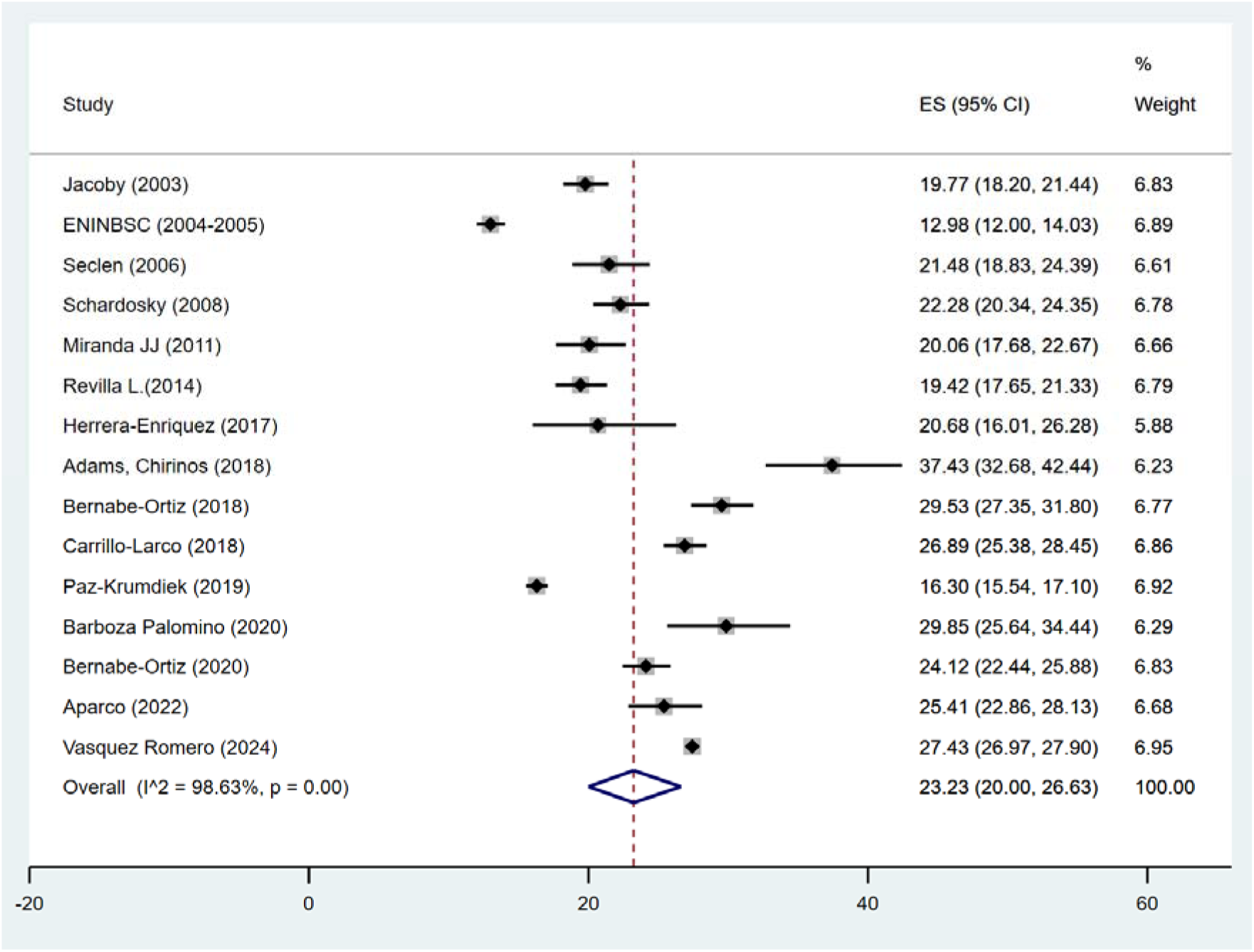
Pooled Prevalence of Obesity According to BMI in Peru Table 3. Sensitivity Analysis of Obesity Prevalence by BMI in Peru

**Table 1.** Summary of Studies on the Prevalence of Obesity (BMI, WC, WHR) in Peru.

| First author, year | City | Inclusion Criteria | Exclusion Criteria | Sample | Gender (% women) | Age (mean, median, or proportion) | Obesity measure evaluated | Diagnostic criteria | Database (year of collection) |
| --- | --- | --- | --- | --- | --- | --- | --- | --- | --- |
| Jacoby, 2003 | Lima, Huánuco, Arequipa, Ucayali | Those who wanted to be part of the study | Those who did not have the cognitive ability to respond to the study | 2322 | 1157 (49.8%) | H: 42.8<br>M: 38.2 | BMI<br>WHtR | $\geq 30$ m/Kg<sup>2</sup><br>men $\geq 94$cm<br>women $\geq 84$cm | JACOBY |
| ENINBSC 2004-2005 MINSA | Nacional | Over 20 years old, oriented in time, space, and place, aware of not having a serious illness | Pregnant, breastfeeding, those who have altered their usual diet due to illness, party and/or celebrations, people with physical disabilities that do not allow normal evaluation of anthropometric indicators, as well as bodybuilders and qualified athletes | 4183 | 2097 (50.13%) | 20 - 38 | BMI<br>WC<br>WHtR | $\geq 30$ m/Kg<sup>2</sup><br>men $\geq 102$ cm<br>women $\geq 88$ cm<br>$\geq 0.50$ | ENINBSC (2004-2005) |
| Seclen, 2006 | Rímac, SMP, Los Olivos | Over 30 years old and residents in the mentioned areas | Type 1 diabetes, pregnant, acute illness, presence of a medical condition that could affect anthropometric measures or laboratory data, illiterate | 838 | 612 (68.3%) | | BMI<br>WC | $\geq 30$ m/Kg<sup>2</sup><br>men $\geq 102$ cm<br>women $\geq 88$ cm | Propia |
| Medina-Lezama, 2007 | Arequipa | Between 20 and 80 years old | Pregnant | 1878 | 1011 (53.83%) | 49.6<br>48.5 | BMI | $\geq 30$ m/Kg<sup>2</sup> | PREVENTION (2005) |
| Schargrodsky, 2008 | Lima | Over 25 years old up to 64 years old | Life limits, pregnant, residents with an address different from their home or residents who lived in areas considered dangerous for interviewers and people who were visiting | 1652 | 883<br>(53.4%) | 43.6 | BMI | ≥ 30<br>m/Kg<sup>2</sup> | CARMELA<br>(2008) |
| Miranda JJ, 2011 | Lima - Ayacucho | Over 30 years old | Those with incomplete laboratories or anthropometric measurements | 987 | 521<br>(52.8%) | 48 | BMI<br>WC<br>WHtR | ≥ 30<br>m/Kg<sup>2</sup><br>men ≥ 102<br>cm<br>women ≥ 88 cm<br>≥ 0.50 | PERU<br>MIGRANT<br>(2007-2008) |
| Revilla L., 2014 | Lima, Callao | Over 15 years old | Under 15 years old, residence less than a year in the study location, pregnant, and disabling conditions that prevent adequate anthropometric evaluation | 1771 | 1105<br>(62%) | 39.5 | BMI<br>WC | ≥ 30<br>m/Kg<sup>2</sup><br>men ≥ 102<br>cm<br>women ≥ 88 cm | FRENT LIMA<br>CALLAO<br>(2007) |
| Herrera-Enriquez, 2017 | Arequipa - Chivay | Individuals over 40 years of age | Individuals with a history or evidence of alcoholism, advanced liver damage with ascites, renal insufficiency, hypothyroidism, Cushing's syndrome, psychiatric illness, pregnant or postpartum women, use of oral contraceptives, hormone replacement therapy, steroids, glucocorticoids, and incomplete clinical history data | 237 | 127<br>(53.6%) | H: 68.9<br>M: 62.0 | BMI<br>WC | ≥ 30<br>m/Kg<sup>2</sup><br>men ≥ 102<br>cm<br>women ≥ 88 cm | Propia |
| Adams, 2018 | Lima | Individuals aged 20 to 59 years, beneficiaries of popular dining halls in Lima | Individuals with terminal chronic disease, mental deficiency or difficulty in expression, consumptive disease, pregnant women, or those with acute illness | 374 | 266<br>(71.1%) |  | BMI<br>WC | ≥ 30<br>m/Kg<sup>2</sup><br>men ≥ 90cm<br>women ≥ 80cm | Propia |
| Bernabé-Ortiz, 2018 | Tumbes | Individuals aged 30 to 69 years, full-time residents in the study area ($\geq 6$ months) and capable of understanding the procedures and providing informed consent | Pregnant women or those with any physical disability that impeded anthropometric measurements (weight, height, blood pressure, or waist circumference) or bedridden individuals. Participants who did not complete laboratory analysis | 1612 | 810<br>(50.3%) | 48.2 | BMI<br>WC | $\geq 30$ m/Kg<sup>2</sup><br>men $\geq 90$cm<br>women $\geq 80$cm | FINDRISCK (2014) |
| Carrillo-Larco, 2018 | Lima, Tumbes, Puno rural y urbano | Individuals over 35 years of age | Pregnant women who could not provide informed consent, individuals incapable of responding to the questionnaires | 3217 | 1660<br>(51.6%) | 55.7 | BMI<br>WC | $\geq 30$ m/Kg<sup>2</sup><br>men $\geq 90$cm<br>women $\geq 80$cm | CRONICAS (2010-2011) |
| Paz-Krumdiek, 2019 | Nacional | Individuals over 30 years of age | Extremes of life, pregnant women, postpartum women, and those not presenting variables of interest | 8587 | 4602<br>(53.6%) | 38.4 | BMI | $\geq 30$ m/Kg<sup>2</sup> | ENAHOCENAN (2011) |
| Barboza Palomino, 2020 | Huamanga (Ayacucho) | Personas entre 18 a 64 años que viven en ayacucho | Pregnant or breastfeeding women, individuals with neurological or mental health issues that impede a logical interview or responses | 412 | | | BMI | $\geq 30$ m/Kg<sup>2</sup> | Propia |
| Bernabé-Ortiz, 2020 | Tumbes | Individuals over 18 years of age, capable of understanding the procedures, able to provide informed consent, and full-time residents in the area | Individuals with self-reported history of chronic kidney disease and heart disease under treatment with digoxin | 2376 | 1197<br>(50.4%) | 43.3 | BMI | $\geq 30$ m/Kg<sup>2</sup> | SALT (2014) |
| Aparco, 2022 | Nacional | Individuals aged 18 to 59 years, fasting for no less than 9 hours and no more than 12 hours | Adults aged 18 to 59 years not on the identification list, pregnant/postpartum women, adults under treatment that alters glucose or lipid profile, those not fasting, individuals with gastrointestinal issues that alter food intake, and genetic conditions or malformations limiting anthropometric techniques | 1047 | 603<br>(57.6%) | 21.4 | BMI<br>WC<br>WHtR | $\geq 30$ m/Kg<sup>2</sup><br>men $\geq 94$ cm<br>women $\geq 88$ cm<br>$\geq 0.50$ | VIANEV (2017-2018) |
| Vasquez Romero, 2024 | Nacional | Individuals aged 20 to 99 years | Pregnant women, absent anthropometric markers, or values outside the normal physiological range | 35 255 | 18 136 (51.44) | 40.78 | BMI<br>WC<br>WHtR | ≥ 30 m/Kg2<br>men ≥ 102 cm<br>women ≥ 88 cm<br>≥ 0.50 | ENDES 2022 |

**Table 2.**
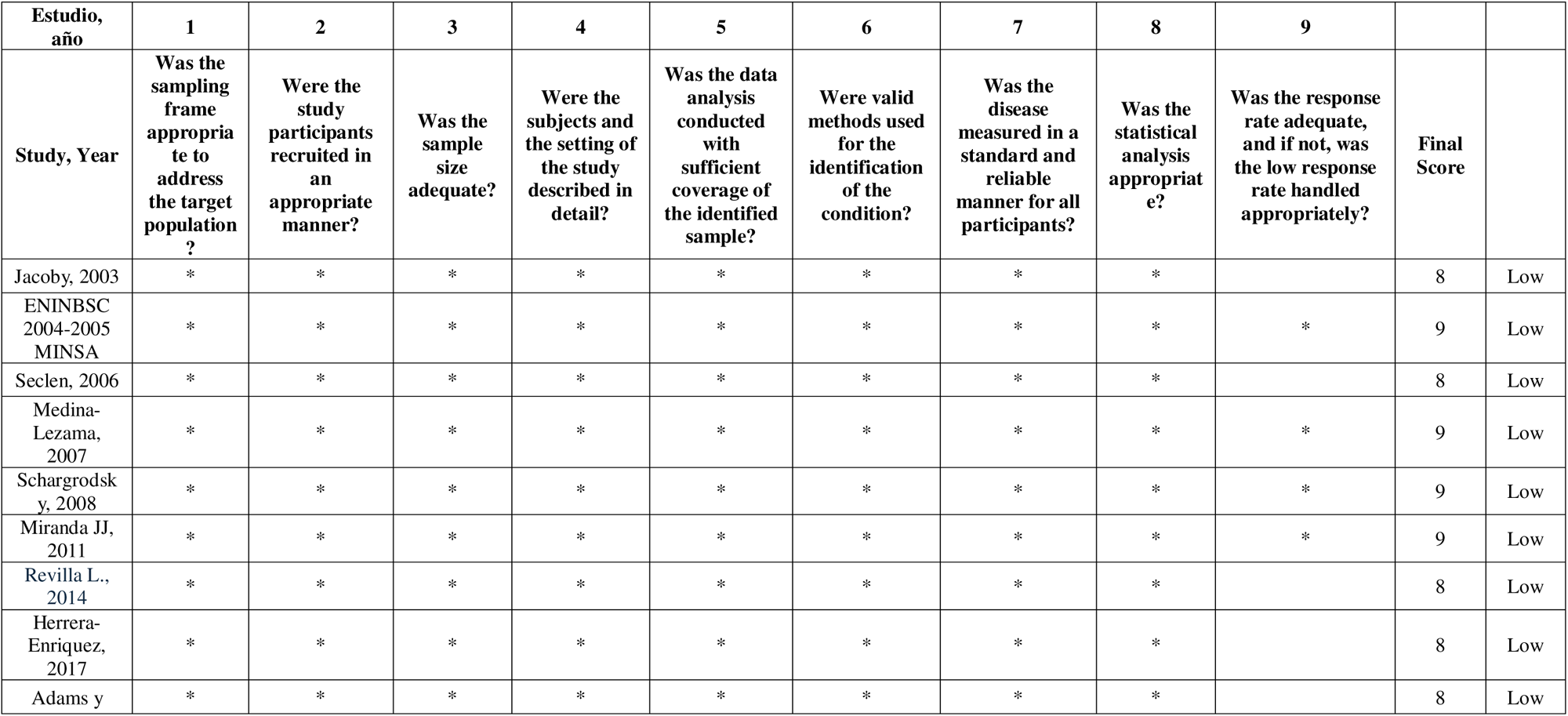
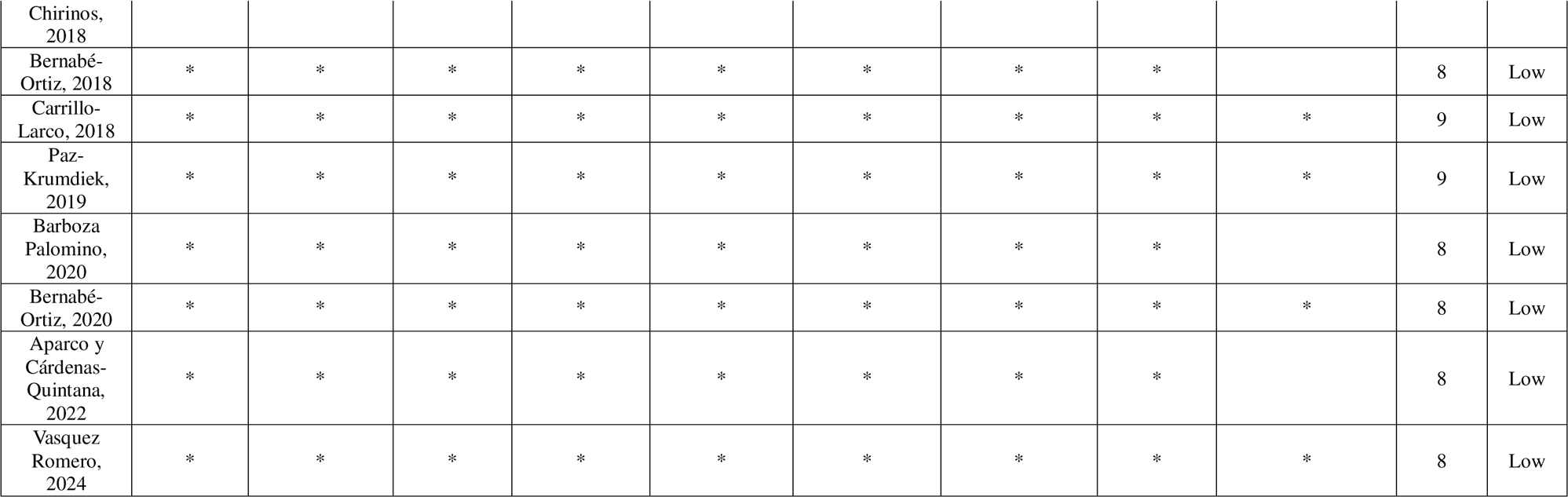
Level of bias in studies on the prevalence of obesity in peru.

**Table 3.** Sensitivity Analysis of Obesity Prevalence by BMI in Peru.

| Category | Number of studies | Prevalence | IC 95% | I<sup>2</sup> |
| --- | --- | --- | --- | --- |
| **Sex** |  |  |  |  |
| Men | 10 | 12.74 | 5.93, 23.22 | 99.09 |
| Women | 10 | 42.9 | 31.33, 54.89 | 99.23 |
| **Time** |  |  |  |  |
| < 2013 | 2 | 29.5 | 28.26, 30.75 |  |
| ≥ 2013 | 2 | 43.11 | 42.61, 43.62 |  |
| **Scope** |  |  |  |  |
| Subnational | 2 | 31.31 | 29.59, 33.05 |  |
| National | 2 | 27.77 | 27.33, 28.21 |  |

The systematic review results indicate significant variations in obesity prevalence measured by BMI across different categories. Ten studies were included for each sex, finding an obesity prevalence in males of 12.74% (95% CI: 5.93% - 23.22%, I² = 99.09%), while in females, the prevalence was considerably higher at 42.9% (95% CI: 31.33% - 54.89%, I² = 99.23%). Analyzing the data over time, with two studies in each category, an increase in obesity prevalence was observed in studies conducted from 2013 onwards (43.11%, 95% CI: 42.61% - 43.62%) compared to those conducted before 2013 (29.5%, 95% CI: 28.26% - 30.75%). Regarding geographical scope, the prevalence in subnational studies was 31.31% (95% CI: 29.59% - 33.05%), while national studies showed a slightly lower prevalence of 27.77% (95% CI: 27.33% - 28.21%).

For obesity, according to WC, only those studies that used cut-off points according to the National Cholesterol Education Program Adult Treatment Panel III (ATP III) were included in the meta-analysis. In this way, the pooled prevalence was 38.90% (95% CI: 37.47% - 46.60%), but the heterogeneity was quite high (I² = 99.02%) (Figure 2).

**Figure 3.**
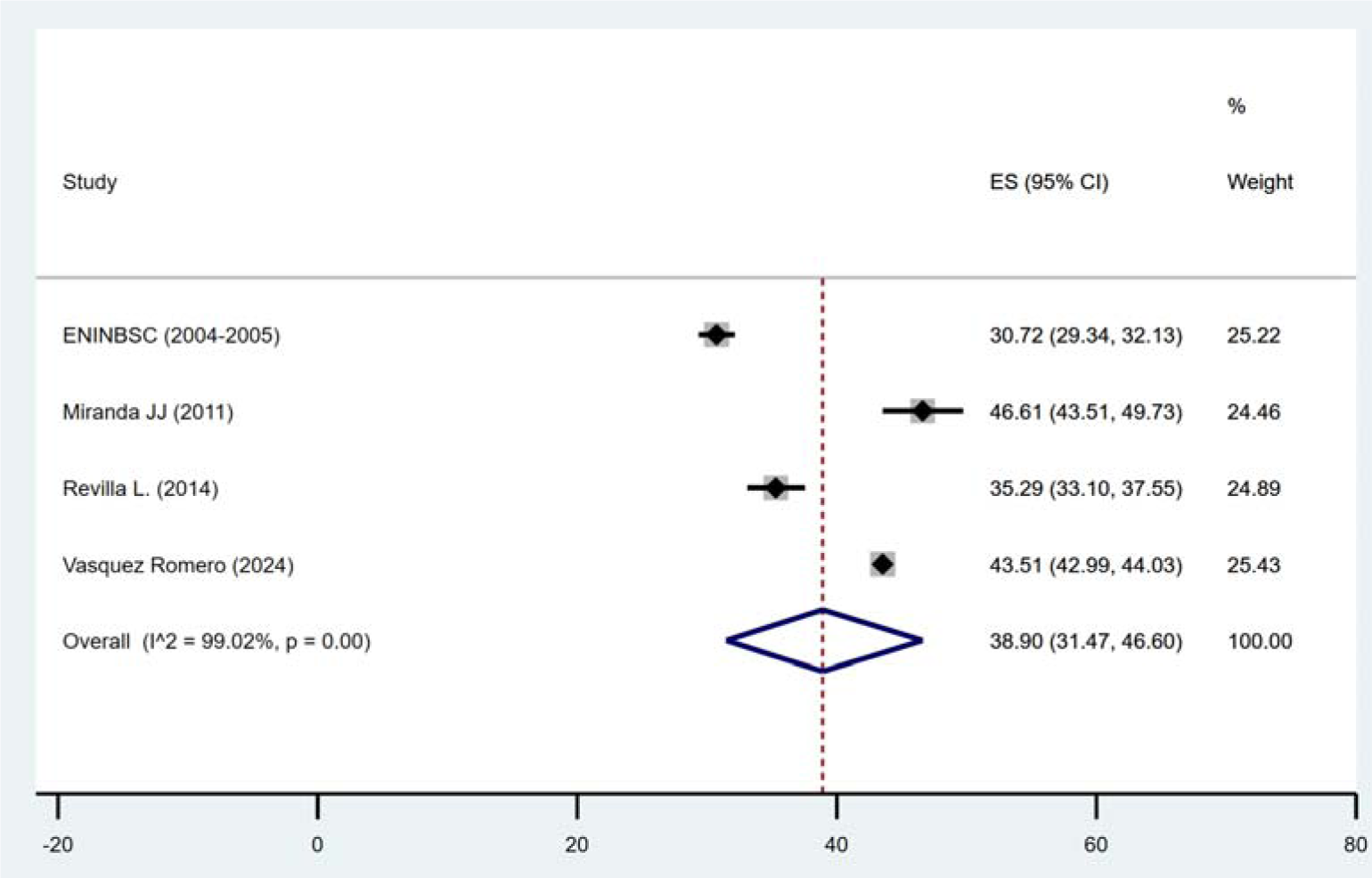
Pooled Prevalence of Obesity According to WC in Peru

### Sensitivity Analysis of Obesity Prevalence by WC in Peru

In the sensitivity analysis of obesity prevalence by WC in Peru, ten studies were evaluated for each sex group. The prevalence of obesity in males was 17.10%, with a 95% confidence interval ranging from 10.03% to 25.58%, showing very high heterogeneity (I² = 98.81%). In contrast, the prevalence in females was significantly higher, estimated at 49.58% (95% CI: 34.87% - 64.32%), with also high heterogeneity (I² = 99.50%). Over time, studies conducted before 2013 reported a prevalence of 33.64% (95% CI: 32.36% - 34.93%), whereas more recent studies conducted from 2013 onwards showed an increase in prevalence to 43.11% (95% CI: 42.61% - 43.62%). From a geographical perspective, subnational studies indicated a prevalence of 39.28% (95% CI: 37.46% - 41.11%), compared to national studies, which reported a lower prevalence of 27.43% (95% CI: 27.33% - 28.21%).

**Table 4.**
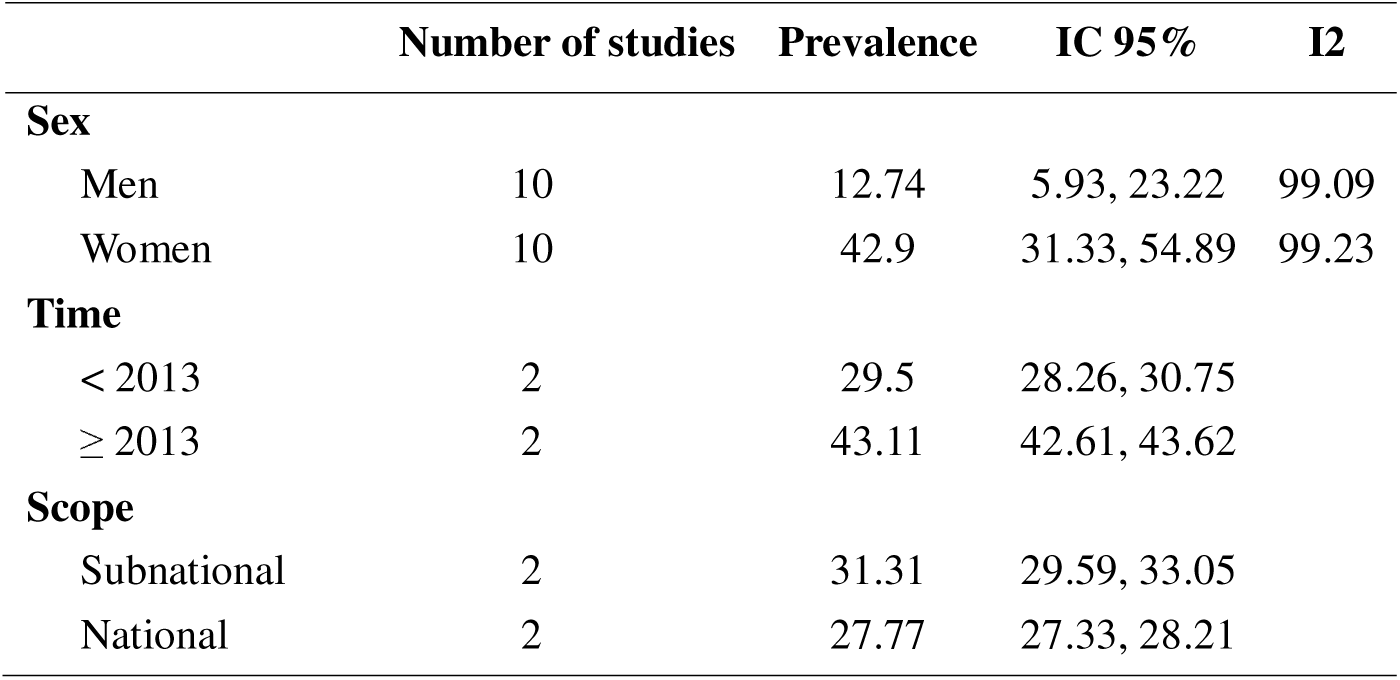
Sensitivity Analysis of Obesity Prevalence by WC in Peru.

According to WHtR, the pooled prevalence for obesity was 81.53% (95% CI: 77.56%— 85.19%), but the heterogeneity was quite high (I² = 98.61%) (Figure 2).

**Figure 4.**
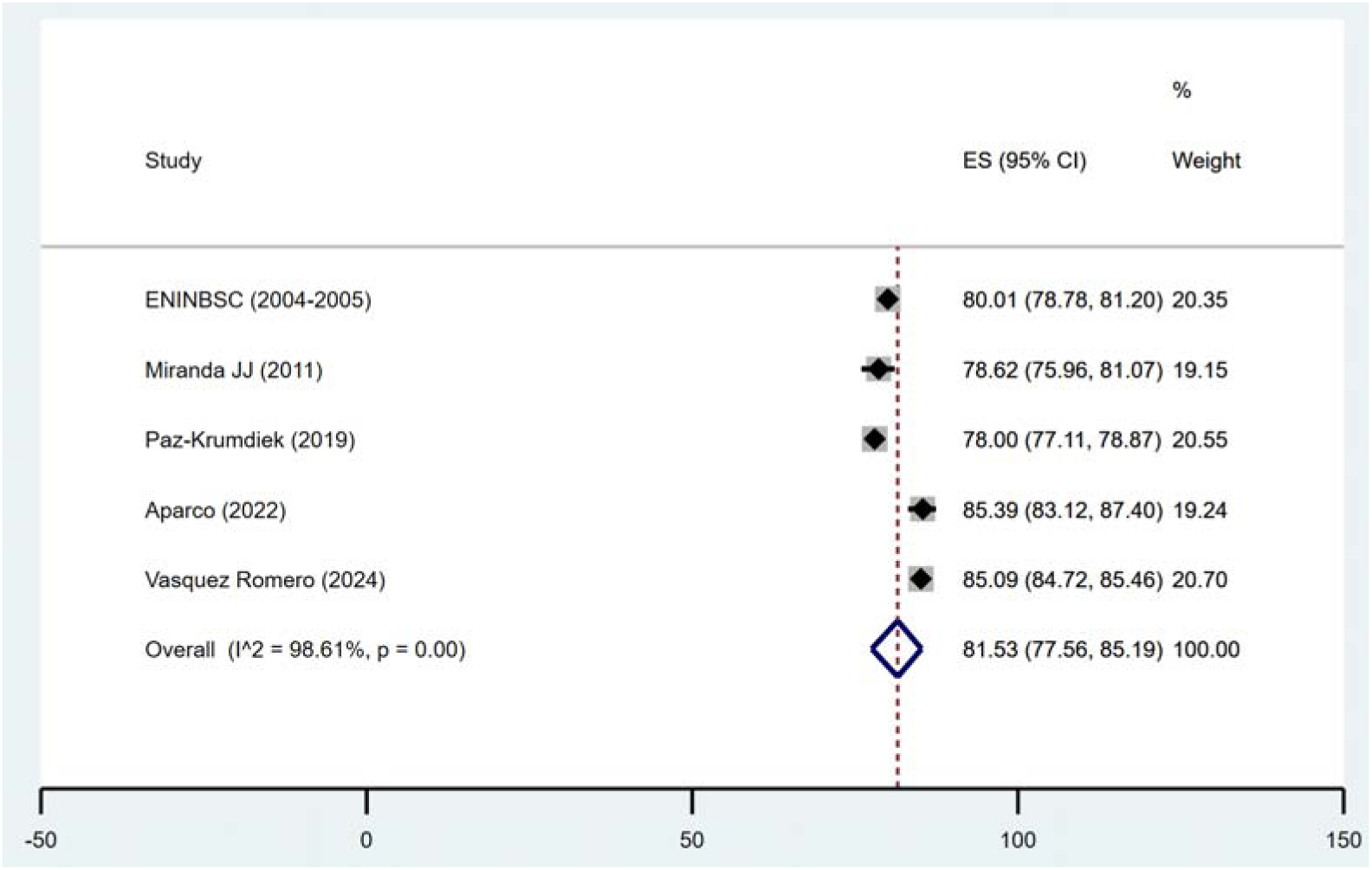
Pooled Prevalence of Obesity According to WHtR in Peru

The sensitivity analysis for obesity prevalence determined by WHtR in Peru included four studies for each sex group. Male prevalence was 76.87% (95% CI: 72.33% - 81.11%), with a heterogeneity of 94.72%. In females, the prevalence was slightly higher, at 87.58% (95% CI: 84.38% - 90.45%), with a heterogeneity of 93.74%. Regarding the time of the studies, those conducted before 2013 presented a prevalence of 76.76% (95% CI: 78.65% - 80.84%), while more recent studies, from 2013 onwards, showed a prevalence of 82.91% (95% CI: 77.33% - 87.85%), with extremely high heterogeneity of 99.15%. Analyzing the geographical scope, subnational studies revealed a prevalence of 82.19% (95% CI: 77.79% - 86.21%), compared to national studies, which showed a similar prevalence of 76.87% (95% CI: 72.33% - 81.11%). Heterogeneity was also very high in both cases, at 98.9% for subnational and 94.72% for national.

**Table 5.**
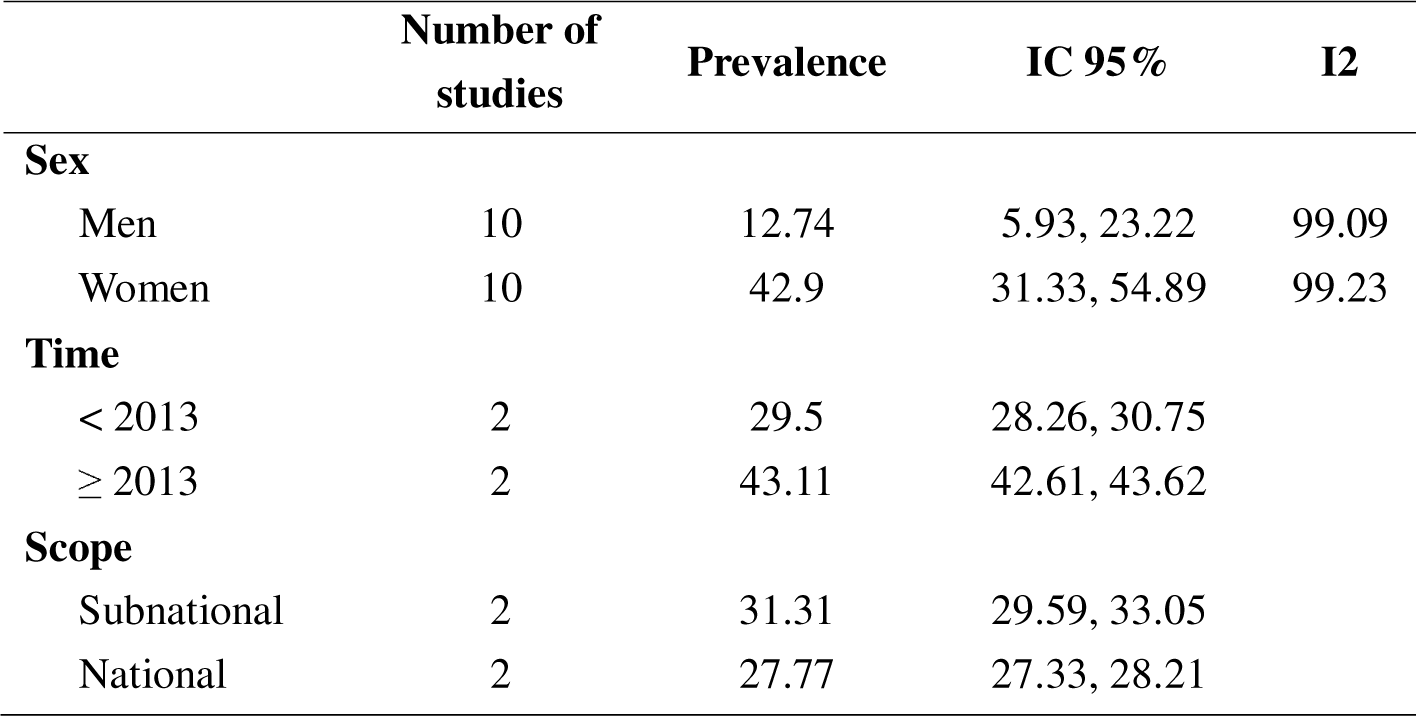
Sensitivity Analysis of Obesity Prevalence by WHtR in Peru.

### Risk of Bias Assessment

The 19 selected articles were evaluated using the risk of bias scale by Munn et al. ^(8)^ for prevalence studies. The evaluated studies showed adequate compliance with criteria to minimize the risk of bias, using appropriate sampling frameworks and effective recruitment methods to select representative samples of the target population. Additionally, all studies confirmed adequate sample sizes, provided detailed descriptions of subjects and settings, and conducted thorough data analyses, ensuring the validity and reliability of their results.

However, the response rate and management were the most variable points. Although not all studies provided explicit details on how a potentially low response rate was handled, those that did maintained a low risk of bias.

## Discussion

This systematic review was designed to assess the prevalence of obesity in Peru using anthropometric parameters such as BMI, WC, and WHtR. Including 16 studies that met rigorous selection criteria, the analysis provides a crucial perspective on the evolution and distribution of obesity across various demographics and regions in the country. However, it is concerning that only five of these studies were published in the last five years ^(4–6,13,20)^, and only two studies ^(5,6)^ aimed to determine obesity prevalence. This indicates a possible lack of recent data that could affect the responsiveness of public health policies to this escalating epidemic.

The results reveal an upward trend in obesity prevalence since 2013, an alarming signal that could be related to lifestyle changes, such as increased physical inactivity and the consumption of processed and high-energy-density foods ^(21)^. Specifically, the gender disparity is notable, with women showing a significantly higher prevalence compared to men. This difference could reflect variations in biological, behavioral, and socioeconomic factors that disproportionately affect women, highlighting the need for public health strategies that consider the particularities of each sex ^(22)^.

The markers used to measure obesity showed considerable variations in their results. WC and WHtR showed prevalences that suggest conventional cut-off points may not be appropriate for the Peruvian population. For example, the ATP III criterion ^(23)^ for WC was preferred over the International Diabetes Federation (IDF) criterion ^(24)^, as the latter estimated a prevalence of 60%, an unrealistic figure for the population under study. Similarly, for WHtR, if the globally consensual cut-off points of ≥ 0.5 ^(25)^ are followed, obesity prevalence would reach up to 80%, a value that, as mentioned earlier, does not reflect reality. Therefore, studies like Vásquez-Romero et al ^(6)^ used the cut-off of 0.59, which provided a more realistic prevalence, although it still yielded equally high values. Considering this, there is a need to review and possibly recalibrate these cut-off points to reflect the true prevalence of obesity in the country more accurately.

### Challenges and Limitations of Current National Surveys

Only four of the 16 studies identified in our systematic review are national in scope, highlighting a significant limitation in the coverage and periodicity of obesity data in Peru. The ENDES survey is conducted annually ^(26)^ but is not specifically designed to measure obesity. Although it includes a section on obesity, its focus is not as comprehensive as a survey dedicated exclusively to this issue. Similarly, the ENAHO ^(27)^ occasionally reports obesity prevalence solely based on BMI, which does not provide a complete picture.

On the other hand, the ENINBSC ^(14)^ and VIANEV ^(28)^ surveys, although more specific, are outdated; the former was conducted in 2005, and the latter in 2017, and they are not updated annually. This contrasts markedly with surveys like Mexico’s ENSANUT ^(29)^, targeted at metabolic problems and updated regularly, providing continuous and up-to-date data crucial for formulating effective public health policies.

This review highlights the urgency of implementing annual national surveys to monitor obesity in all dimensions, including BMI and WC. Greater investment in such studies would allow for more accurate and representative data on the prevalence of obesity in Peru, facilitate monitoring trends over time, and assess the effectiveness of implemented interventions.

Establishing annual and specific surveys for obesity would help align Peru with international standards and enable a more agile and informed response to this growing public health challenge. These studies should include various anthropometric markers to provide a more comprehensive view of the population’s nutritional and health status. Additionally, they should be designed to capture significant regional and demographic differences, allowing for more targeted and effective interventions.

The Peruvian government and public health agencies must recognize the importance of these data and provide the necessary resources for their systematic collection and analysis. Updating and expanding the data infrastructure on obesity would facilitate a better understanding of the epidemic, the adaptation of public policies to changing realities, and the implementation of more effective prevention and treatment strategies based on solid and up-to-date evidence.

### Importance of the Study for Public Health

The importance of this study for public health in Peru is significant, given the growing obesity epidemic and its associated consequences. Obesity is not just an individual health problem but a public health challenge that incurs high costs for the healthcare system, reduces quality of life, and increases mortality. By providing an updated assessment of obesity prevalence using various anthropometric markers, this study offers a crucial foundation for understanding the scale of the problem and identifying the population groups at higher risk. This information is vital for designing and implementing effective and targeted public health policies and intervention programs.

Moreover, the study emphasizes the need to improve data collection and the frequency of health surveys related to obesity in Peru. The lack of recent and specific data on obesity limits the ability of the government and health organizations to develop policies that effectively address this crisis. With more frequent and specific studies, such as annual and nationally representative surveys, policymakers would have access to information that more accurately reflects current trends and the effectiveness of ongoing interventions. This is essential for assessing the progression of obesity at the national level and adjusting intervention strategies as new evidence emerges.

Finally, this study highlights the importance of adjusting the cut-off points and methodologies used to measure obesity in the Peruvian context, which could result in more accurate estimates and, consequently, more appropriate interventions. Adapting international standards to local characteristics is crucial for accurately assessing the population’s health and developing health programs that address Peru’s cultural, economic, and social particularities. Ultimately, conducting such studies strengthens the country’s ability to manage public health more effectively, offering hope for a significant reduction in the burden of obesity for future generations.

### Study Limitations

A crucial limitation of this study is the limited availability of recent research specifically focused on obesity. Another significant limitation is the variability in methods and cut-off points used to define obesity among the different studies included in the review. Differences in how obesity is defined and measured, such as the cut-off points for WC and WHtR, can lead to discrepancies in reported prevalences. This lack of standardization complicates direct comparisons between studies and may affect the accuracy of national prevalence estimates, hindering the formulation of effective intervention strategies.

Moreover, although some included studies are national in scope, many are subnational and may not fully capture regional variations within Peru. This limitation is relevant because it prevents identifying specific areas or population groups that require urgent interventions. Additionally, the infrequency of national studies reduces the ability to continuously assess the impact of public health interventions and adjust policies according to emerging needs.

## Conclusions

This study has provided a comprehensive view of the prevalence of obesity in Peru using a variety of anthropometric markers. Overall, the results reveal an obesity prevalence of 23.23%, 38.90%, and 81.53% according to BMI, WC, and WHtR, respectively, with increasing trends disproportionately affecting women. The review also highlighted significant variations in measurement methodologies and cut-off points used to determine obesity, leading to discrepancies in reported prevalences. Furthermore, the insufficiency of recent and specific data on obesity, especially in national studies, limits the ability to effectively track and adapt public health policies in response to this epidemic.

It is recommended that the collection and frequency of data on obesity in Peru be improved by implementing annual national surveys specifically designed for this purpose. These surveys should include a variety of anthropometric markers, such as weight, height, and WC, to obtain a more accurate and comprehensive representation of the epidemic. Additionally, it is crucial to standardize measurement methodologies and review the cut-off points used to adapt them to the specific characteristics of the Peruvian population, thereby improving the accuracy of prevalence estimates and allowing for more effective comparisons over time and between different studies.

From a public health policy perspective, emphasis should be placed on developing strategies that not only focus on reducing the prevalence of obesity but also address its root causes. This includes implementing educational programs on nutrition, promoting physical activity, and ensuring access to healthy foods. Policies and programs should be inclusive, considering gender and regional differences, and culturally appropriate to ensure effectiveness. Moreover, continuous research should be encouraged to monitor trends and evaluate the effectiveness of implemented interventions.

## Acknowledgments

We want to express my gratitude to the members of the Institute of Tropical Disease Research, Universidad Nacional Toribio Rodríguez de Mendoza de Amazonas (UNTRM), Amazonas, Peru, for their support and contributions throughout the conduct of this research.

## Informed Consent

Informed consent was not required for this study.

## Data Availability

Data are available upon request from the corresponding author.

## Author Contributions

Luisa Erika Milagros Vásquez-Romero: Conceptualization, Investigation, Methodology, Resources, Writing - Original Draft, Writing - Review & Editing

Fiorella E. Zuzunaga-Montoya: Investigation, Project administration, Writing - Original Draft, Writing - Review & Editing

Joan A. Loayza-Castro: Investigation, Resources, Writing - Original Draft, Writing - Review & Editing

Enrique Vigil-Ventura: Validation, Visualization, Writing - Original Draft, Writing - Review & Editing

Willy Ramos: Software, Data Curation, Formal analysis, Writing - Review & Editing

Víctor Juan Vera-Ponce: Methodology, Supervision, Funding acquisition, Writing - Review & Editing

## Supplementary file 1

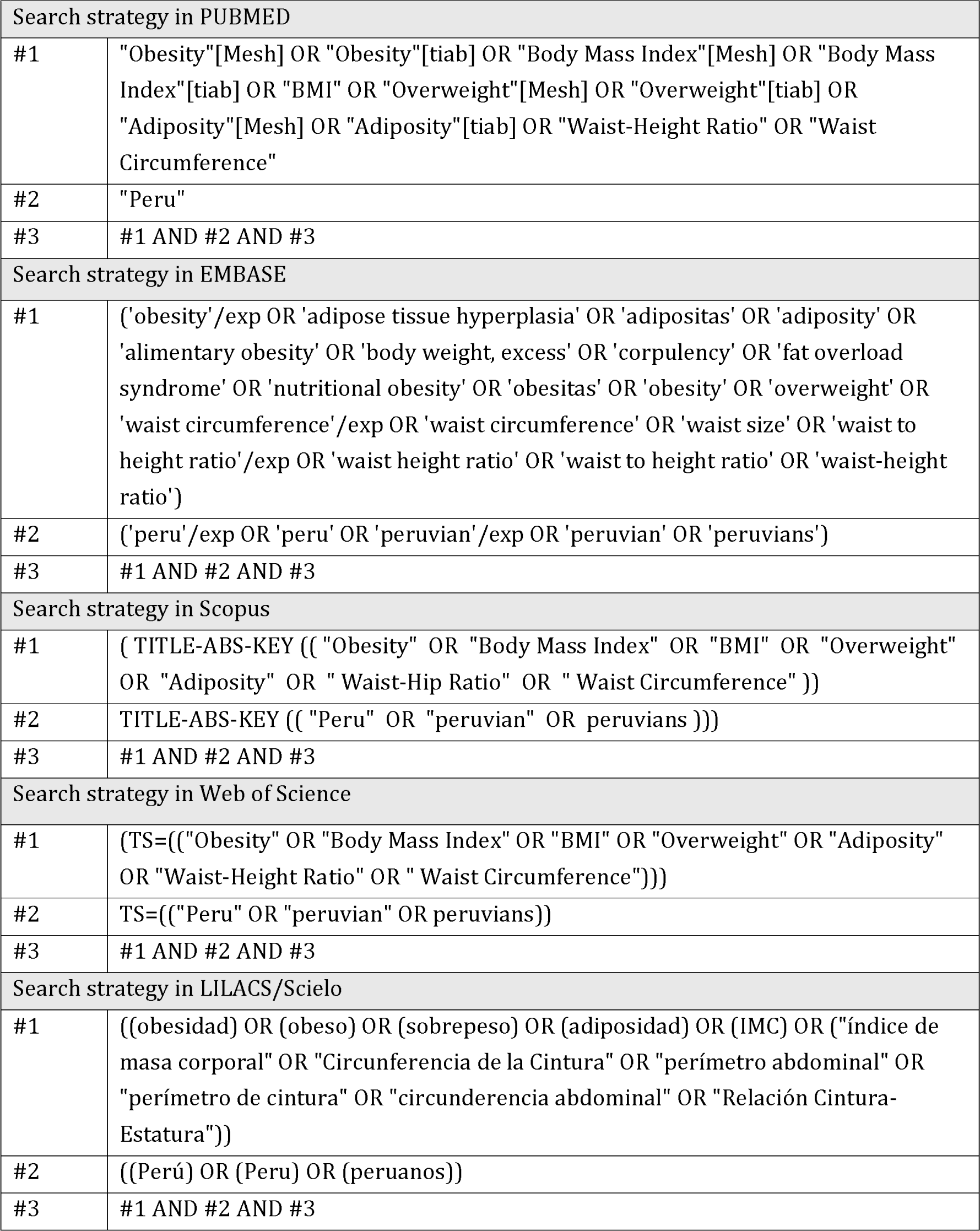

## References

1. Obesity and overweight [Internet]. [cited August 18, 2023]. Available at: https://www.who.int/news-room/fact-sheets/detail/obesity-and-overweight

2. Organización Mundial de la Salud. Documento de debate de la OMS: Proyectos de Recomendaciones para la prevención y el tratamiento de la obesidad a lo largo del curso de la vida, incluidas las posibles metas. OMS. 2021;1–12.

3. Kovalskys I, Fisberg M, Gómez G, Pareja RG, Yépez García MC, Cortés Sanabria LY, et al. Energy intake and food sources of eight Latin American countries: results from the Latin American Study of Nutrition and Health (ELANS). Public Health Nutr. 2018;21(14):2535–47. doi:10.1017/S1368980018001222

4. Paz-Krumdiek M, Rodriguez-Vélez SG, Mayta-Tristán P, Bernabe-Ortiz A. Association between sitting time and obesity: A population-based study in Peru. Nutr Diet. 2020;77(2):189–95. doi:10.1111/1747-0080.12540

5. Aparco JP, Cárdenas-Quintana H. Correlación y concordancia del índice de masa corporal con el perímetro abdominal y el índice cintura-talla en adultos peruanos de 18 a 59 años. Revista Peruana de Medicina Experimental y Salud Publica. 2022;39(4):392–9. doi:10.17843/rpmesp.2022.394.11932

6. Romero LEMV, Vera-Ponce VJ, Zuzunaga-Montoya F, Torres-Malca JR, Loayza-Castro J, Paucar CRI, et al. Associated factors, concordance, and trends of obesity: body mass index, abdominal waist, and waist- to-height ratio between 2014 and 2022. Analysis of nine national survey. 2024. doi:10.21203/rs.3.rs-3745026/v2

7. Page MJ, McKenzie JE, Bossuyt PM, Boutron I, Hoffmann TC, Mulrow CD, et al. The PRISMA 2020 statement: an updated guideline for reporting systematic reviews. BMJ. 2021;n71. doi:10.1136/bmj.n71

8. Munn Z, Moola S, Lisy K, Riitano D, Tufanaru C. Methodological guidance for systematic reviews of observational epidemiological studies reporting prevalence and cumulative incidence data. Int J Evid Based Healthc. 2015;13(3):147–53. doi:10.1097/XEB.0000000000000054

9. Jacoby E, Goldstein J, López A, Núñez E, López T. Social class, family, and life-style factors associated with overweight and obesity among adults in Peruvian cities. Prev Med. 2003;37(5):396–405. doi:10.1016/s0091-7435(03)00159-2

10. Herrera-Enriquez K, Narvaez-Guerra O. Discordance of metabolic syndrome and abdominal obesity prevalence according to different criteria in Andean highlanders: A community-based study. Diabetes Metab Syndr. 2017;11 Suppl 1:S359–64. doi:10.1016/j.dsx.2017.03.016

11. Seclén S, Villena A, Larrad MT, Gamarra D, Herrera B, Pérez CF, et al. Prevalence of the metabolic syndrome in the mestizo population of peru. Metab Syndr Relat Disord. 2006;4(1):1–6. doi:10.1089/met.2006.4.1

12. Adams KJ, Chirinos JL. Prevalence of Risk Factors for Metabolic Syndrome and Its Components in Community Kitchen Users in a District in Lima, Peru. Rev Peru Med Exp Salud Publica. 2018;35(1):39–45. doi:10.17843/rpmesp.2018.351.3598

13. Palomino EEB. Prevalencia de factores de riesgo para enfermedades crónicas no transmisibles en Perú. Revista Cuidarte. 2020;11(2). doi:10.15649/cuidarte.1066

14. Instituto Nacional de Salud. Encuesta Nacional de Indicadores Nutricionales, Bioquimicos, Socioeconomicos y Culturales relacionados con las Enfermedades Cronicas Degenerativas. Lima, Peru: Instituto Nacional de Salud: Centro Nacional de Alimentacion y Nutricion. 2006.

15. Medina-Lezama J, Zea-Diaz H, Morey-Vargas OL, Bolaños-Salazar JF, Muñoz-Atahualpa E, Postigo-MacDowall M, et al. Prevalence of the metabolic syndrome in Peruvian Andean hispanics: the PREVENCION study. Diabetes Res Clin Pract. 2007;78(2):270–81. doi:10.1016/j.diabres.2007.04.004

16. Revilla L, López T, Sánchez S, Yasuda M, Sanjinés G. Prevalencia de hipertensión arterial y diabetes en habitantes de Lima y Callao, Perú. Revista Peruana de Medicina Experimental y Salud Publica. 2014;31(3):437–44.

17. Carrillo-Larco RM, Miranda JJ, Gilman RH, Checkley W, Smeeth L, Bernabé-Ortiz A, et al. Trajectories of body mass index and waist circumference in four Peruvian settings at different level of urbanisation: the CRONICAS Cohort Study. J Epidemiol Community Health. 2018;72(5):397–403. doi:10.1136/jech-2017-209795

18. PERU MIGRANT Study | Baseline dataset [Internet]. figshare; 2016 [citado el 14 de marzo de 2021]. doi:10.6084/m9.figshare.3125005.v1

19. Miranda JJ, Gilman RH, García HH, Smeeth L. The effect on cardiovascular risk factors of migration from rural to urban areas in Peru: PERU MIGRANT Study. BMC Cardiovasc Disord. 2009;9:23. doi:10.1186/1471-2261-9-23

20. Bernabe-Ortiz A, Sal y Rosas VG, Ponce-Lucero V, Cárdenas MK, Carrillo-Larco RM, Diez-Canseco F, et al. Effect of salt substitution on community-wide blood pressure and hypertension incidence. Nat Med. 2020;26(3):374–8. doi:10.1038/s41591-020-0754-2

21. Vera Ponce VJ, Torres Malca JR, Tello Quispe EK, Orihuela Manrique EJ, De La Cruz Vargas JA. Validación de escala de cambios en los estilos de vida durante el periodo de cuarentena en una población de estudiantes universitarios de Lima, Perú. Rev Fac Med Hum. 2020;614–23.

22. Garawi F, Devries K, Thorogood N, Uauy R. Global differences between women and men in the prevalence of obesity: is there an association with gender inequality? Eur J Clin Nutr. 2014;68(10):1101–6. doi:10.1038/ejcn.2014.86

23. Expert Panel on Detection, Evaluation, and Treatment of High Blood Cholesterol in Adults. Executive Summary of The Third Report of The National Cholesterol Education Program (NCEP) Expert Panel on Detection, Evaluation, And Treatment of High Blood Cholesterol In Adults (Adult Treatment Panel III). JAMA. 2001;285(19):2486–97. doi:10.1001/jama.285.19.2486

24. International Diabetes Federation. IDF Diabetes Atlas [Internet]. 10th ed. Brussels: International Diabetes Federation; 2021 [cited 2024 Feb 14]. Available from: https://diabetesatlas.org/idfawp/resource-files/2021/07/IDF_Atlas_10th_Edition_2021.pdf.

25. Browning LM, Hsieh SD, Ashwell M. A systematic review of waist-to-height ratio as a screening tool for the prediction of cardiovascular disease and diabetes: 0·5 could be a suitable global boundary value. Nutr Res Rev. 2010;23(2):247–69. doi:10.1017/S0954422410000144

26. Perú: Encuesta Demográfica y de Salud Familiar - ENDES 2022 [Internet]. [citado el 1 de marzo de 2024]. Disponible en: https://www.gob.pe/institucion/inei/informes-publicaciones/4233597-peru-encuesta-demografica-y-de-salud-familiar-endes-2022

27. Encuesta Nacional de Hogares (ENAHO) 2022 - [Instituto Nacional de Estadística e Informática – INEI] | Plataforma Nacional de Datos Abiertos [Internet]. [citado el 4 de marzo de 2024]. Disponible en: https://www.datosabiertos.gob.pe/dataset/encuesta-nacional-de-hogares-enaho-2022-instituto-nacional-de-estad%C3%ADstica-e-inform%C3%A1tica-%E2%80%93

28. Centro Nacional de Alimentación y Nutrición. Estado Nutricional En Adultos de 18 a 59 Años VIANEV 2017–2018. Instituto Nacional de Salud: Lima, Peru. 2021;191.

29. Encuesta Nacional de Salud y Nutrición [Internet]. ENCUESTAS. [citado el 2 de mayo de 2024]. Disponible en: https://ensanut.insp.mx/index.php

